# A Deep Learning Approach to Accurately Discriminate Between Optic Disc Drusen and Papilledema on Fundus Photographs

**DOI:** 10.1101/2023.05.24.23290447

**Authors:** Kanchalika Sathianvichitr, Raymond P. Najjar, Tang Zhiqun, J. Alexander Fraser, Christine Wen Leng Yau, Michael Julien Alexandre Girard, Fiona Costello, Mung Yan Lin, Wolf Alexander Lagrèze, Catherine Vignal-Clermont, Clare L. Fraser, Steffen Hamann, Nancy J. Newman, Valérie Biousse, Dan Milea, the BONSAI Group

## Abstract

**Objective:** To assess the performance of a deep learning system (DLS) to discriminate between optic disc drusen (ODD) and papilledema caused by intracranial hypertension, using standard color ocular fundus photographs collected in a large international multi-ethnic population.

**Design:** Retrospective study.

**Participants:** The study included 4,508 color fundus images in 2,180 patients from 30 neuro-ophthalmology centers (19 countries) participating in the Brain and Optic Nerve Study with Artificial Intelligence (BONSAI) Group.

**Methods:** We trained, validated, and tested a dedicated DLS for binary classification of ODD vs. papilledema (including various subgroups within each category), on conventional mydriatic digital ocular fundus photographs. For training and internal validation, we used 857 ODD images and 3,230 papilledema images, in 1,959 patients. External-testing was subsequently performed on an independent dataset (221 patients) including 207 images with ODD (96 visible and 111 buried), provided by 3 centers of the Optic Disc Drusen Studies Consortium, and 214 images of papilledema (92 mild-to-moderate and 122 severe) from a previously validated study.

**Main outcome measures:** Area under the receiver operating characteristic curve (AUC), accuracy, sensitivity, and specificity were used to discriminate between ODD and papilledema.

**Results:** Overall, the DLS could accurately distinguish between all ODD and papilledema (all severities included): AUC 0.97 (95% confidence interval [CI], 0.96 to 0.98), accuracy 90.5% (95% CI, 88.0% to 92.9%), sensitivity 86.0% (95% CI, 82.1% to 90.1%), and specificity 94.9% (95% CI, 92.3% to 97.6%). The performance of the DLS remained high for discrimination of buried ODD from mild-to-moderate papilledema: AUC 0.93 (95% CI, 0.90 to 0.96), accuracy 84.2% (95% CI, 80.2%-88.6%), sensitivity 78.4% (95% CI, 72.2% to 84.7%), and specificity 91.3% (95% CI, 87.0% to 96.4%).

**Conclusions:** A dedicated DLS can accurately distinguish between ODD and papilledema caused by elevated intracranial pressure, even when considering buried ODD vs mild-to-moderate papilledema. Future studies are required to validate the utility of this DLS in clinical practice.

## Introduction

Papilledema (optic disc swelling from intracranial hypertension) is an important ocular fundus finding, visible on funduscopic examination or standard retinal photographic images. Its clinical identification is not always easy, especially when attempted by non-ophthalmic healthcare providers (neurologists, general practitioners, etc.).^1^ Detection of papilledema prompts urgent critical investigations (brain imaging, cerebrospinal fluid examination, etc.) in order to identify potentially life-or vision-threatening conditions; failure to detect papilledema in a timely manner can lead to visual loss and irreversible neurological sequelae.

Other optic nerve head abnormalities can mimic papilledema (i.e., pseudopapilledema), leading sometimes to unnecessary, invasive and costly investigations.^2,3^ Optic disc drusen (ODD) represent a common cause of misdiagnosed papilledema, especially if the ODD are not visible on funduscopic examination, so-called “buried” ODD. Accurate diagnosis of ODD, which are present in 1.0-2.0% of the general population,^4-6^ is important in clinical practice, not only to discriminate ODD from papilledema, but also because of the possible visual complications from ODD.^7^ Diagnosing ODD is clinically straightforward in the presence of visible, calcified lesions. However, discriminating ODD from true papilledema can be challenging, especially when the optic disc elevation is moderate. Several ocular imaging modalities are helpful in this context, including B-scan orbital ultrasonography,^8,9^ fundus autofluorescence, and more recently, spectral domain enhanced depth imaging optical coherence tomography (OCT).^10-12^ Indeed, OCT has become a valued investigation for ODD diagnosis, although it requires expertise to facilitate image interpretation. Recently, the application of deep learning (DL) methods to OCT imaging has improved the performance of OCT in distinguishing ODD from papilledema, an approach which requires further validation.^13^

Standard color photography of the ocular fundus is an easily accessible modality that can be implemented in non-ophthalmic environments,^14,15^ and is potentially useful for discriminating ODD and papilledema.^16,17^ Fundus photography has recently regained new interest for detection of optic nerve disease, with some successful applications of DL for accurate diagnosis.^18^ A large international study group (Brain and Optic Nerve Study with Artificial Intelligence, BONSAI) showed that a DLS could detect papilledema and determine its severity on color fundus images, with a performance which was comparable to that of expert neuro-ophthalmologists.^19-21^ The main aim of the initial BONSAI study was to distinguish papilledema from normal discs, and from a group of “other” optic disc abnormalities, including a mix of ODD, optic atrophy, and anterior ischemic optic neuropathies. Although this classification can be very helpful as the initial approach in a patient with optic disc abnormalities, there is an additional clinical need to discriminate accurately between pseudopapilledema (buried ODD being its main cause) and true papilledema, because the vision and life-threatening implications for the patients may be radically different.

The aim of our current study was to develop, train and test a new dedicated DLS, able to specifically discriminate between ODD and true papilledema on standard color ocular fundus images. In addition, we evaluated the classification performance of the DLS with various clinically relevant subclasses of papilledema severity and ODD visibility.

## Methods

### Standard Protocol Approvals, Registrations, and Patient Consents

The study was approved by the Centralized Institutional Review Board of SingHealth, Singapore as well as by ethical committees of each contributing institution, and was conducted in accordance with the Declaration of Helsinki. Informed consent was exempted, given the retrospective nature of the study and the use of deidentified medical information and ocular fundus images.

### Study Population

The primary training and internal validation dataset consisted of 4,087 ocular fundus images, among which there were 857 images of optic discs from patients with confirmed ODD and 3,230 images of optic discs with confirmed papilledema, retrospectively collected from 1,959 patients from 30 participating centers of BONSAI (Table 1).

The external testing dataset consisted of 221 patients and 421 images (207 with ODD and 214 with papilledema). The ODD images were provided by three independent expert centers participating in the Optic Disc Drusen Studies (ODDS) Consortium^10,22^: 1/ University of Calgary (Alberta, Canada), 2/ Rigshospitalet, University of Copenhagen (Denmark) and 3/ Western University (Ontario, Canada). Images with papilledema were selected at random from four participating centers: 1/ Ramathibodi Hospital, Mahidol University (Thailand), 2/ University of Freiburg (Germany), 3/ Farabi Eye Hospital, Tehran University of Medical Sciences (Iran), and 4/ Angers University Hospital (France), as previously described elsewhere.^21^ All the images included in the external-testing dataset were independent from the images used in the training dataset.

We excluded patients with concurrent ophthalmic pathologies (e.g., ODD co-existing with optic disc swelling or ODD co-existing with other optic nerve or retinal disease). We also excluded images with poor quality or with poorly centered optic discs. Thus, among the 4,574 initially available images, we excluded 66 (1.4%) images (Fig 1).

### Image Acquisition

Fundus images and relevant clinical information were retrospectively collected in a mix of consecutive and convenience samples in multiple international neuro-ophthalmology centers. Fundus images were obtained in eyes with pharmacologically dilated pupils, using various commercial desktop digital fundus cameras (Table S1, Supplemental Material) by neuro-ophthalmologists who routinely obtain fundus images and who have access to the patients’ medical records. The fundus images were centered on either the macula or the optic disc at various fields of view (subtending 20° to 45°). Fundus images obtained by wide-field fundus camera or with multicolor imaging mode were excluded from this study. Deidentified unaltered fundus images were transferred to the Singapore Eye Research Institute for further analysis.

### Classification of ODD

Patients with ODD were enrolled by the expert neuro-ophthalmology providers only if the diagnosis was confirmed by at least one of the following imaging modalities^10^: 1/ spectral domain enhanced depth imaging optical coherence tomography (EDI-OCT) of the optic nerve head, in agreement with the ODDS Consortium acquisition protocol^22^; 2/ swept-source OCT (SS-OCT) of the optic nerve head; 3/ other ODD diagnostic methods, including B-scan orbital ultrasonography, and fundus autofluorescence.

After diagnosis confirmation, ODD were further classified into “visible ODD” and “buried ODD.” Visible ODD were defined by the presence of visually identifiable refractile bodies on the optic disc surface; the remaining confirmed ODD by ancillary investigations were classified as “buried.”^23-25^ In the primary training dataset, the classification of “visible” vs “buried” ODD was made post-hoc, individually, by two neuro-ophthalmologists at the Singapore Eye Research Institute, on the 857 available color fundus images, using a dedicated semi-automated image presentation software.^26^ A senior neuro-ophthalmologist adjudicated the 41(4.8%) images with discordant diagnosis to obtain consensus.

In the external-testing dataset, the classification “visible” vs “buried” was provided directly by the local neuro-ophthalmology experts from the three centers participating in the ODDS Consortium, based on a combination of the appearance of the ODD on the color fundus images and the results of ancillary imaging investigations (most often, EDI-OCT).

### Classification of papilledema severity

Papilledema images were obtained in patients with confirmed intracranial hypertension due to 1/ a known secondary cause (i.e. intracranial mass, hydrocephalus, cerebral venous thrombosis, medication, etc.) or 2/ idiopathic intracranial hypertension, according to the modified Dandy criteria and abnormally elevated cerebrospinal fluid opening pressure on lumbar puncture.^27^ These images were further classified according to the papilledema severity by two expert neuro-ophthalmologists into two classes, as previously described.^21^ In brief, the standard Frisén 5-grade severity classification was simplified into a 2-grade classification: (1) mild-to-moderate papilledema, corresponding to Frisén grades 1-3, and (2) severe papilledema, corresponding to Frisén grades 4 and 5. The images with discordant severity were adjudicated by 2 additional neuro-ophthalmologists and a consensus was obtained for all images.

### Development of the Deep-Learning Classification Model

#### Data splitting

The primary dataset was randomly split, according to common practices for deep learning model development, into training/validation (80%) and internal-testing datasets (20%). A stratified split was also applied to reduce technical bias among centers. Among the 4,087 images used for this purpose, 3,357 images (82.1%) were used for training/validation, and 730 images (17.9%) were used for the internal evaluation of the model. A stratified split was also used to divide the training/validation dataset. Eventually, 2,856 of 3,357 images (85.1%) were used for training the model and 501 (14.9%) for validation or model parameter tuning.

#### Image Segmentation

The segmentation network was based on U-Net architecture with a ResNet-34 encoder,^28^ using an image resolution of 256×256 pixels, to identify the optic disc and peripapillary region (region of interest, ROI). The automatically segmented ROIs were then resized to 456×456 pixels as the input images for the classification network.

#### Image Preprocessing and Sampling

After image segmentation, we used data augmentation techniques including random rotation, horizontal, warp, zoom, translation shifts, and random drop out of certain input regions.^29^ Oversampling was applied to the minority class (ODD images).

#### Model Training

We used a classification network (EfficientNet-B5), initialized using weights pretrained on ImageNet^30^ and fine-tuned in an end-to-end manner to achieve the best performance. The EfficientNet-B5 architecture was chosen due to its high efficiency for image classification, while using fewer resources and computing power.^31^ The systematically compound scale-up algorithm composed of mobile inverted bottleneck convolution (MBConv) blocks (Fig 2) has improved performance compared to other DLS.^19^ Label smoothing was used to minimize the cross-entropy loss function. The classification network was trained and tuned on 3,357 fundus images (training and validation dataset) to automatically classify the ROIs into specific classes of discs with ODD or discs with papilledema.

#### Model Selection, performance evaluation and heatmaps generation

After selecting the best predictive model (based on preliminary evaluation of the internal-testing dataset), we evaluated its performance on 421 fundus images from the independent external-testing dataset. We first assessed the model’s performance for discriminating between all ODD images (with both visible and buried ODD) and all papilledema images (irrespective of their severity). Then, we evaluated more specifically the performance of the same DLS for tasks with clinically increasing difficulty: 1/ discrimination between visible ODD and severe papilledema, 2/ discrimination between visible ODD and mild-to-moderate papilledema, 3/discrimination between buried ODD and severe papilledema, 4/ discrimination between buried ODD and mild-to-moderate papilledema.

Finally, we generated heatmaps using the class-specific gradient information (Gradient-weighted Class Activation Mapping, Grad-CAM) extracted from the final convolutional layer of the CNN model to visualize the relative relevance of pixels in the input images for the classification task.^32^

### Statistical Analysis

To evaluate the performance of the DLS to distinguish between discs with ODD and discs with papilledema (and their respective subclasses), we calculated various metrics, including the area under the receiver operating characteristic curve (AUC), sensitivity, specificity, and accuracy. Bootstrapping (2,000 times) with patient as the sampling unit was used to estimate 95% confidence intervals (CI) of the performance metrics.

## Results

### Patient and Image Characteristics

For training and internal validation purposes, the study included 4,087 images, collected from 1959 patients (91.2% with images of both eyes, 9.9% with images on follow-up visits) recruited by 30 sites participating in the BONSAI consortium for developing the model. We included 857 ODD images and 3,230 papilledema images. The external-testing dataset included images from seven international sites. In total, 421 fundus images from 221 patients (90.5% with images of both eyes, none of the images duplicative of the same eye) were enrolled. Among them, 207 ODD images (111 buried ODD and 96 visible ODD, Table 2) were collected from three sites participating in the ODDS Consortium; and 214 papilledema images (92 images of discs with mild-to-moderate papilledema and 122 images of discs with severe papilledema), which were previously validated as described above, were included.

Patient demographics and image characteristics are described in Table 2. Altogether, the ratio between visible/buried drusen varied according to the age of patients, with higher proportions of buried drusen at lower ages. In the training/validation group, the frequency of buried ODD was 33.3% in patients over 40, reaching 96.9% in patients under 11; in the external-testing dataset, the frequency of buried ODD in those age groups was 44.3% and 83.3%, respectively (Table 3).

### Overall Classification Performance

In the validation, internal-testing, and external-testing datasets, the DLS was able to discriminate discs with confirmed ODD from discs with papilledema with AUCs of 0.99 (95% CI, 0.98 to 1.00), 0.98 (95% CI, 0.97 to 0.99), and 0.97 (95% CI, 0.96 to 0.98) (Table 4). In the internal-testing dataset, the DLS achieved a sensitivity of 91.9% (95% CI, 90.1% to 94.3%) and specificity of 93.9% (95% CI, 90.1% to 97.3%) for an overall accuracy of 93.2% (95% CI, 91.1% to 94.7%). In the external-testing dataset, the DLS had an overall sensitivity of 86.0% (95% CI, 82.1% to 90.1%), a specificity of 94.9% (95% CI, 92.3% to 97.6%), and an accuracy of 90.5% (95% CI, 88.0% to 92.9%). Examples of representative heatmaps allowing for the visualization of pixels with the most discriminative value for classification tasks by the DLS are shown in Fig 3.

### Subclass performance (buried/visible ODD versus mild-moderate/severe papilledema)

Next, we assessed the performance of the DLS in the external-testing datasets, specifically for the two subgroups of ODD (visible and buried) and the two classes of papilledema severity (Table 5). The DLS performance followed a gradient, with the highest parameters for distinguishing visible ODD and severe papilledema (AUC 0.99 [95% CI, 0.98 to 1.00], accuracy 96.3% [95% CI, 94.4% to 98.6%]), followed by marginally decreased performance for distinguishing visible ODD from mild-to-moderate papilledema (AUC 0.96 [95% CI, 0.94 to 0.98], accuracy 93.1% [95% CI, 90.2% to 96.1%]). The performance of discriminating buried ODD from papilledema at all stages showed only slightly reduced performance (AUC 0.96 [95% CI, 0.95 to 0.98], accuracy 89.2% [95% CI, 86.4% to 92.2%]). Unsurprisingly, the performance of the DLS was excellent in discriminating buried ODD from severe papilledema (AUC 0.99 [95% CI, 0.98 to 1.00], accuracy 88.4% [95% CI, 85.0% to 92.1%]). The lowest performance of the DLS was noted for distinguishing buried ODD from mild-to-moderate papilledema (AUC 0.93 [95% CI, 0.90 to 0.96], accuracy 84.2% [95% CI, 80.2% to 88.6%]) (Fig 4).

### Post-hoc analysis of classification errors

Forty retinal fundus images (9.5% of the 421 images) in the external-testing dataset were wrongly diagnosed by the DLS compared to the ground truth. More specifically, the DLS misclassified 11 papilledema images as ODD, in 5.1% of the total of 214 papilledema images. The majority of these misclassified images represented mild papilledema (n=8, 72.7%). Among the 207 images of confirmed ODD, 29 images (14.0%) were wrongly diagnosed as papilledema. The large majority of the misdiagnosed ODD were buried ODD (24 images, 82.8%).

## Discussion

The main finding of this study is that a trained DLS achieved excellent performance at discriminating ODD from true papilledema on standard color ocular fundus images, without any additional clinical information. The performance of the DLS across various subgroups of ODD and papilledema images followed a decreasing trend which was compatible with the reality of everyday clinical practice. Indeed, just as clinicians do in real life, the DLS displayed a higher performance in discriminating visible ODD from severe papilledema. Conversely, the performance of the DLS dropped (but only marginally) in the most difficult clinical situation of mild elevation of the optic disc (typically seen in buried ODD and mild-to-moderate papilledema).

Accurate distinction between ODD and papilledema is crucial to avoid unnecessary, invasive, and costly procedures or, conversely, to detect vision-or life-threatening conditions. Among several ancillary investigations aiming to discriminate between ODD and papilledema, fluorescein angiography has been reported to have high accuracy and good interobserver agreement,^12,33^ but it remains an invasive procedure, reducing its applicability outside ophthalmic departments. Fundus autofluorescence is a cost-effective, non-invasive method requiring little subjective interpretation for detecting superficial ODD. However, it doesn’t reliably detect ODD located in the deeper portions of the optic nerve head.^34^ Ultrasonography was until recently considered the most accurate investigation for diagnosing ODD, and, more generally, for distinguishing true papilledema from pseudopapilledema.^35^ However, it is less able to detect poorly calcified ODD or buried ODD,^36^ and is also a highly operator-dependent procedure, difficult to implement outside dedicated clinics.

OCT is commonly used for ODD diagnosis and is considered as gold standard by the international ODDS Consortium.^22^ The advent of EDI-OCT has facilitated visualization of deep structures in the optic nerve head, including ODD that are deeply located underneath Bruch’s membrane opening. In discs with papilledema, transverse axial and en face OCT reveal structural changes, including anterior displacement of Bruch’s membrane opening, peripapillary folds or wrinkles, and peripapillary fluid accumulation.^37-39^ Quantitative OCT measures of the peripapillary RNFL thickness also may be helpful in the evaluation and monitoring of the optic disc swelling of papilledema.^40-43^ However, there are several limitations in clinical use, including the considerable overlap of RNFL thickness values among normal optic discs, discs with deep ODD or mild papilledema,^44,45^ absence of normative RNFL thickness values in children and in ocular conditions such as high myopia, falsely normative values of RNFL thickness when there is concurrent optic disc edema and optic atrophy, and false positively thickened RNFL from other disc abnormalities (e.g., gliosis, hyperopia).^46^

In 2020, the BONSAI Study Group reported that an artificial intelligence-based DLS can be successfully applied to standard ocular fundus color photograph for the detection of papilledema with an excellent AUC of 0.96, a sensitivity of 96.4%, and a specificity of 84.7%.^19^ However, that DLS did not address specifically the clinically relevant discrimination between entities that can mimic each other such as true papilledema and ODD. For this purpose, we subsequently developed a next-generation DLS, after inclusion of further robust imaging and clinical data provided by international expert neuro-ophthalmologists. The model was developed using a recent advanced deep convolutional neural network, EfficientNet-B5, which already has been successfully applied in the fields of radiology^47^ and, more recently, in ophthalmology to detect vision-threatening diabetic retinopathy.^48^

The advantages of our study are its considerably large and multiethnic population, which is, to our knowledge, the largest studied population comparing papilledema with ODD. We also believe that the ground truth for ODD and papilledema in this study was robustly accurate. This is particularly true for patients with papilledema, as the presence of elevated intracranial pressure was confirmed in every case, either by results of brain imaging or by the cerebrospinal fluid opening pressure. Furthermore, we have confidence in our results regarding the diagnosis of ODD and its subgroups given that: 1) the distribution of the ODD (visible vs buried) according to age groups was comparable in the training and the testing datasets, and also aligned with previous findings in the literature^49^; indeed, as in our study (Table 3), the frequency of visible ODD increases with age; and 2) the three referring centers of ODD images are members of the international ODDS Consortium, a group of clinicians with a specific interest and expertise in the detection and study of ODD.

Regarding limitations, this is a retrospective study based on convenience samples, acquired by expert neuro-ophthalmologists who provided high-quality images obtained with mydriatic desktop cameras; therefore, we cannot yet know if these results can be extrapolated to images obtained with non-mydriatic or handheld cameras. Additionally, we have not assessed patients with coexistent optic disc pathologies (i.e., ODD and papilledema), although clinically, it is likely a rare occurrence.

**In conclusion**, our DLS applied to color ocular fundus images accurately discriminates between eyes with true papilledema and eyes with ODD at all severities; its performance decreases only marginally when challenged to make the clinically difficult distinction between eyes with buried ODD and eyes with mild-to-moderate papilledema. Further prospective studies are needed to confirm the applicability of such a DLS in real clinical settings.

## Supporting information

The BONSAI Consortium Collaborator List

## Data Availability

All data produced in the present study are available upon reasonable request to the authors

## Acknowledgements

we thank Megan Tay Mei Chen and Jodi Ling Wei Yan for their precious assistance during this study.

