## Supplementary material for "A Deep Learning Approach to Accurately Discriminate Between Optic Disc Drusen and Papilledema on Fundus Photographs": Figures 1-4 and Tables 1-5

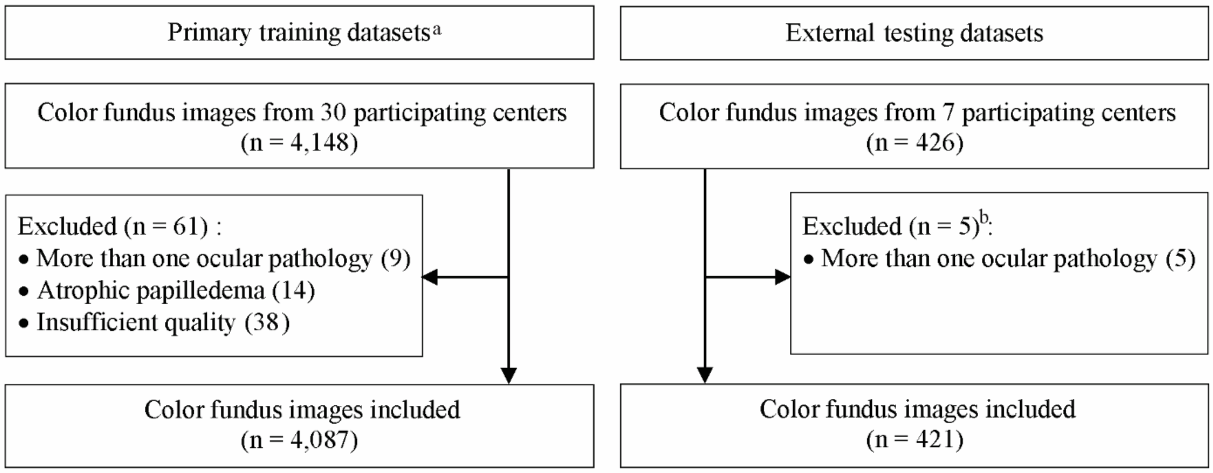


**Fig 1. Flowchart displaying the process of inclusion of fundus photographs in the training and testing datasets.**

[a] Primary training datasets included training, validation, and internal-testing datasets.

[b] Details are provided only for the optic disc drusen, the details regarding papilledema photographs are provided elsewhere.[21]

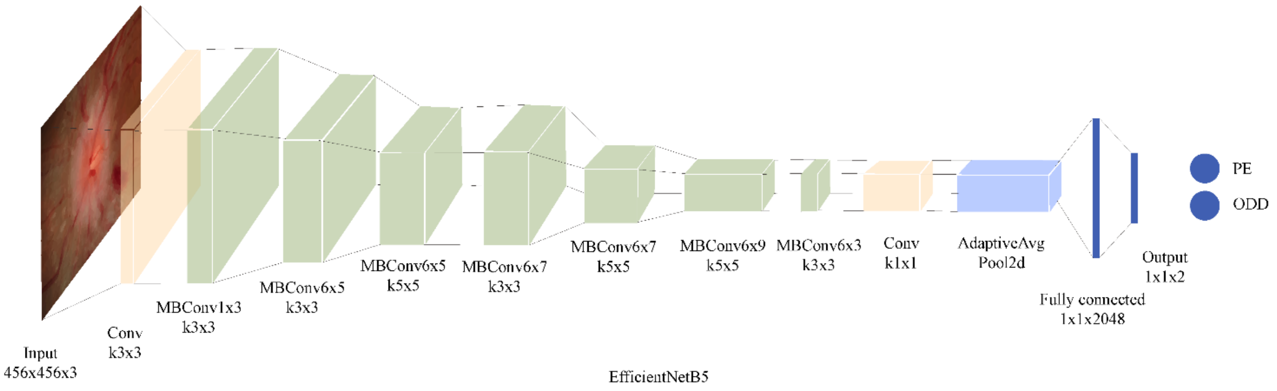


**Fig 2. Deep learning classification network.**

Abbreviation: AdaptiveAvg Pool2d = 2D adaptive average pooling; Conv = convolution; MBConv = mobile inverted bottleneck convolution; ODD = optic disc drusen; PE = papilledema.

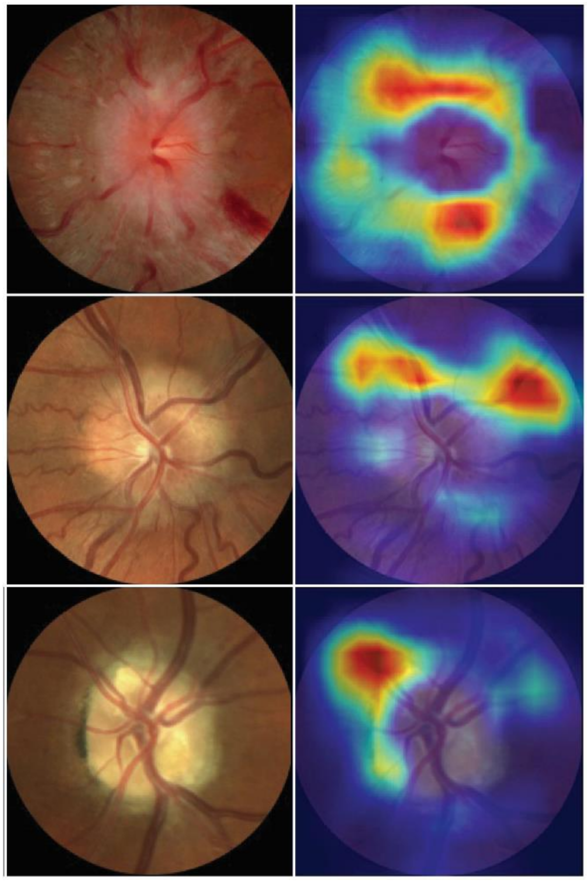

**Fig 3**. **Examples of color fundus photographs centered on the optic disc and corresponding class activation maps (heatmaps).**

Upper row: Optic disc photograph, accurately predicted as “papilledema” by the algorithm. The heatmap displays a circular, ring-shaped activation, at the margins of the optic disc. Middle row: optic disc photograph of confirmed buried optic disc drusen, accurately diagnosed by the algorithm (probability of ODD = 96.74% vs. probability of papilledema = 3.26%). Bottom row: photograph of an optic disc with visible optic disc drusen which was accurately identified by the algorithm (probability = 99.99%). The activation region (right column) displays a region that is superimposed on the visible drusen (left column, 11 o’clock).

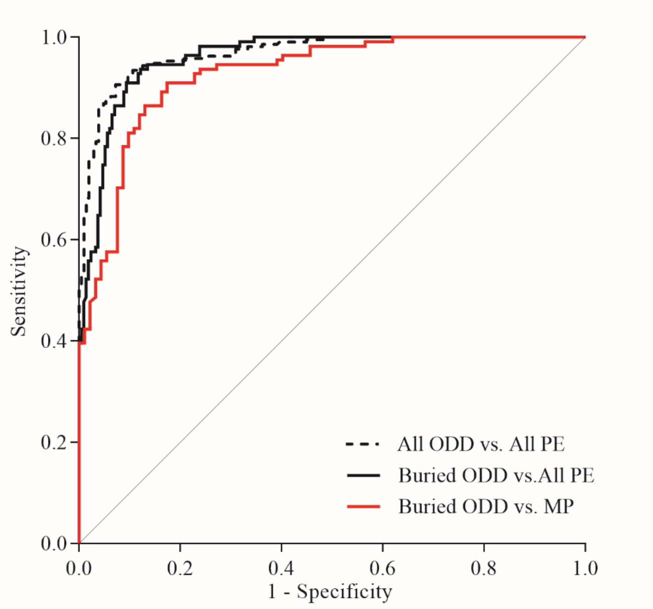


**Fig 4. Performance of the deep learning system for discriminating between buried optic disc drusen (ODD) and papilledema.**

The deep learning system discriminated ODD (both visible and buried) from papilledema (at all stages of severity), with an area under the receiver operating characteristic curve (AUC) of 0.97 (95% CI, 0.96 to 0.98). The system could discriminate between buried ODD and papilledema (at all stages of severity) with an AUC of 0.96 (95% CI, 0.95 to 0.98) and between buried ODD from discs with mild-to-moderate papilledema (AUC 0.93 [95% CI, 0.90 to 0.96]).

**Table 1** List of Participating Centers and Number of Images Included per Center

| Participating center | Discs with ODD | Discs with PE | Total |
|---|---|---|---|
| | No. of images | | |
| **Training, validation, and internal testing datasets** | | | |
| **Angers, France** | 34 | 307 | 341 |
| **Atlanta, GA, United States** | 316 | 1231 | 1547 |
| **Baltimore, MD, United States** | 37 | 134 | 171 |
| **Bangkok, Thailand** | 3 | 0 | 3 |
| **Bologna, Italy** | 11 | 13 | 24 |
| **Bordeaux, France** | 21 | 45 | 66 |
| **Chennai, India** | 53 | 293 | 346 |
| **Calgary, Alberta, Canada** | 0 | 100 | 100 |
| **Coimbra, Portugal** | 18 | 24 | 42 |
| **Copenhagen, Denmark** | 23 | 92 | 115 |
| **Freiburg, Germany** | 88 | 61 | 149 |
| **Geneva, Switzerland** | 4 | 14 | 18 |
| **Grenoble, France** | 8 | 6 | 14 |
| **Guangzhou, China** | 2 | 0 | 2 |
| **Hongkong, China** | 8 | 16 | 24 |
| **Kinshasa, Democratic Republic of Congo** | 0 | 8 | 8 |
| **London, Ontario, Canada** | 0 | 276 | 276 |
| **London, United Kingdom** | 20 | 34 | 54 |
| **Manila, Philippines** | 2 | 11 | 13 |
| **Paris, France** | 2 | 101 | 103 |
| **Rochester, MN, United Sates** | 21 | 96 | 117 |
| **Seoul, Korea** | 107 | 129 | 236 |
| **Singapore, Singapore** | 2 | 72 | 74 |
| **Sydney, Australia** | 24 | 50 | 74 |
| **Syracuse, NY, United States** | 16 | 22 | 38 |
| **Tehran, Iran** | 25 | 5 | 30 |
| **Toronto, Ontario, Canada** | 8 | 86 | 94 |
| **Ufa, Russia** | 4 | 4 | 8 |
| **Total Training, validation, and internal testing datasets** | 857 | 3230 | 4087 |
| **External-testing datasets** | | | |
| **Angers, France** | 0 | 10 | 10 |
| **Bangkok, Thailand** | 0 | 36 | 36 |
| **Calgary, Alberta, Canada** | 24 | 0 | 24 |
| **Copenhagen, Denmark** | 101 | 0 | 101 |
| **Freiburg, Germany** | 0 | 86 | 86 |
| **London, Ontario, Canada** | 82 | 0 | 82 |
| **Tehran, Iran** | 0 | 82 | 82 |
| **Total external-testing data sets** | 207 | 214 | 421 |

Abbreviations: ODD = optic disc drusen; PE = papilledema.

**Table 2** Demographics and Image Characteristics in the Training, Validation and Testing Datasets

| Characteristics | Training and validation | Internal testing | External testing | Total |
|---|---|---|---|---|
| **Number of patients (images)** | 1606 (3357) | 353 (730) | 221 (421) | 2180 (4508) |
| **Age, y, mean (SD)** | 33.0 (15.3) [a] | 30.3 (14.7)[b] | 33.8 (16.9) [c] | 33.0 (15.4) |
| **Female, n (%)** | 1189 (77.8) [a] | 263 (78.3)[b] | 158 (71.5) [c] | 1610 (77.2) |
| **Images with buried ODD, n (%)**[d] | 403 (56.5) | 97 (67.4) | 111 (53.6) | |
| **Images with visible ODD, n (%)**[d] | 310 (43.5) | 47 (32.6) | 96 (46.4) | |

[a] On the basis of 95.2% available patient demographic data.
[b] On the basis of 95.2% available patient demographic data.
[c] On the basis of 100.0% available patient demographic data.
[d] Percent referred to the total number of all ODD images in each column.

**Table 3** The Frequency Distribution of Buried Optic Disc Drusen Stratified by Age Groups

| Age groups | Training and Validation [a] | | Internal testing [b] | | External testing [c] | |
|---|---|---|---|---|---|---|
| | **No. of all ODD images** | **No. of buried ODD (%)** | **No. of all ODD images** | **No. of buried ODD (%)** | **No. of all ODD images** | **No. of buried ODD (%)** |
| <11 | 65 | 63 (96.9) | 20 | 20 (100.0) | 6 | 5 (83.3) |
| 11-20 | 184 | 139 (75.5) | 37 | 33 (89.2) | 51 | 36 (70.6) |
| 21-30 | 90 | 55 (61.1) | 25 | 16 (64.0) | 45 | 25 (55.6) |
| 31-40 | 80 | 44 (55.0) | 19 | 10 (52.6) | 35 | 14 (40.0) |
| >40 | 261 | 87 (33.3) | 32 | 9 (28.1) | 70 | 31 (44.3) |
| Total | 680 | 388 (57.1) | 133 | 88 (66.2) | 207 | 111 (53.6) |

[a] On the basis of 95.4% available age data
[b] On the basis of 92.4% available age data
[c] On the basis of 100.0% available age data

**Table 4** Classification Performance of the Deep-Learning System in Discriminating Optic Disc Drusen from Papilledema in the Validation, Internal-Testing Data Set and External-Testing Data Set

| | Total | ODD | PE | AUC | Sensitivity, % | Specificity, % | Accuracy, % |
|---|---|---|---|---|---|---|---|
| | No. of images | | | (95% CI) | | | |
| **Validation dataset** | | | | | | | |
| **ODD vs. PE** | 501 | 105 | 396 | 0.99 (0.98-1.00) | 93.3 (89.5-97.1) | 97.2 (95.8-98.8) | 96.4 (95.0-97.8) |
| **Internal-testing dataset** | | | | | | | |
| **ODD vs. PE** | 730 | 144 | 586 | 0.98 (0.97-0.99) | 91.9 (90.1-94.3) | 93.9 (90.1-97.3) | 93.2 (91.1-94.7)) |
| **External-testing dataset** | | | | | | | |
| **ODD vs. PE** | 421 | 207 | 214 | 0.97 (0.96-0.98) | 86.0 (82.1-90.1) | 94.9 (92.3-97.6) | 90.5 (88.0-92.9) |

Abbreviations: AUC = area under the receiver-operating-characteristic curve; CI = confidence interval; ODD = optic disc drusen; PE = papilledema.

**Table 5** Classification Performance of the Deep-Learning System in Discriminating Different Subgroups of Optic Disc Drusen and Papilledema in the External-Testing Data Set

| | **Total** | **ODD** | **PE** | **AUC** | **Sensitivity, %** | **Specificity, %** | **Accuracy, %** |
|---|---|---|---|---|---|---|---|
| | No. of images | | | (95% CI) | | | |
| **Visible ODD vs. SP** | 218 | 96 | 122 | 0.99 (0.98-1.00) | 94.8 (91.7-98.4) | 97.5 (95.7-100.0) | 96.3 (94.4-98.6) |
| **Visible ODD vs. all PE** | 310 | 96 | 214 | 0.98 (0.97-0.99) | 94.8 (91.5-98.4) | 94.9 (92.4-97.7) | 94.8 (92.8-97.0) |
| **Visible ODD vs. MP** | 188 | 96 | 92 | 0.96 (0.94-0.98) | 94.8 (91.8-98.4) | 91.3 (86.5-96.4) | 93.1 (90.2-96.1) |
| **Buried ODD vs. SP** | 233 | 111 | 122 | 0.99 (0.98-1.00) | 78.4 (72.1-84.9) | 97.5 (95.6-100.0) | 88.4 (85.0-92.1) |
| **Buried ODD vs. all PE** | 325 | 111 | 214 | 0.96 (0.95-0.98) | 78.4 (72.2-85.0) | 94.9 (92.3-97.7) | 89.2 (86.4-92.2) |
| **Buried ODD vs. MP** | 203 | 111 | 92 | 0.93 (0.90-0.96) | 78.4 (72.2-84.7) | 91.3 (87.0-96.4) | 84.2 (80.2-88.6) |

Abbreviations: AUC = area under the receiver-operating-characteristic curve; CI = confidence interval; MP = mild-to-moderate papilledema; ODD = optic disc drusen; PE = papilledema; SP = severe papilledema.
