## Supplemental Table S1 for "A Deep Learning Approach to Accurately Discriminate Between Optic Disc Drusen and Papilledema on Fundus Photographs"

**Table S1** Cameras Used at Participating Centers

| City, country | Camera brand | Model |
|---|---|---|
| Angers, France | Topcon | TRC-NW6S |
| Atlanta, GA, United States | Topcon | TRC-50DX |
| Baltimore, MD, United States | Carl Zeiss | FF4 |
| Bangkok, Thailand | Kowa | WX3D |
| Bologna, Italy | Carl Zeiss/Topcon | VISUCAM 500/DRI OCT Triton |
| Bordeaux, France | Carl Zeiss | VISUCAM 500 |
| Chennai, India | Carl Zeiss | FF450 Plus IR |
| Calgary, Alberta, Canada | Carl Zeiss | VISUCAM 224/524 |
| Coimbra, Portugal | Topcon | TRC-NW7SF Mark II |
| Copenhagen, Denmark | Topcon | TRC-50DX and TRC-NW8 |
| Freiburg, Germany | Carl Zeiss | SF420 |
| Geneva, Switzerland | Carl Zeiss | FF450 Plus |
| Grenoble, France | Topcon/Canon | TRC-NW6S/CR2 |
| Guangzhou, China | Topcon/Carl Zeiss | TRC-50DX/FF450 Plus IR |
| Hongkong, China | Topcon | TRC-50DX |
| Kinshasa, Democratic Republic of Congo | Carl Zeiss | VISUCAM 500 |
| London, Ontario, Canada | Topcon | TRC-50DX |
| London, United Kingdom | Topcon/Canon | TRC-50DX/CR2 |
| Manila, Philippines | Carl Zeiss/ Meditec | VISUCAM 500/VISUCAM NMFA |
| Paris, France | Canon | CRDGI |
| Rochester, MN, United Sates | Topcon | TRC-50DX |
| Seoul, Korea | Kowa | VX-10a |
| Singapore, Singapore | Topcon/ Canon/ Kowa | TRC-50DX/ DRI OCT Triton Plus/ CR-Dgi/ Nonmyd-WX3D |
| Sydney, Australia | Carl Zeiss | VISUCAM 500 |
| Syracuse, NY, United States | Topcon/Carl Zeiss | TRC-NW8 and TRC-NW400/FF 450 |
| Tehran, Iran | Canon | CR2 |
| Toronto, Ontario, Canada | Carl Zeiss | VISUCAM 500 |
| Ufa, Russia | Carl Zeiss | VISUCAM 500 |
