## Supplementary material for "A Deep Learning Approach to Accurately Discriminate Between Optic Disc Drusen and Papilledema on Fundus Photographs": The BONSAI Consortium Collaborator List

### Collaborators of the BONSAI group for this study and their Affiliations (alphabetical order by city)

| Name | Centre |
|---|---|
| Axel Petzold | Neuro-ophthalmology Expert Centre, Amsterdam University Medical Center, Amsterdam, Netherlands |
| Maillette de Buy Wenniger L.J. | Neuro-ophthalmology Expert Centre, Amsterdam University Medical Center, Amsterdam, Netherlands |
| Philippe Gohier | University Hospital Angers, Angers, France |
| Neil R Miller | Johns Hopkins University School of Medicine, Baltimore, MD, USA<br>Division of Neuro-Ophthalmology, Wilmer Eye Institute, Johns Hopkins University, Baltimore, MD, USA |
| Kavin Vanikieti | Faculty of Medicine Ramathibodi Hospital, Mahidol University, Bangkok, Thailand |
| Yanin Suwan | Faculty of Medicine Ramathibodi Hospital, Mahidol University, Bangkok, Thailand |
| Giulia Amore | Dipartimento di Scienze Biomediche e Neuromotorie, Università degli Studi di Bologna, Bologna, Italy |
| Valerio Carelli | IRCCS Istituto delle Scienze Neurologiche di Bologna, UOC Clinica Neurologica, Dipartimento di Scienze Biomediche e Neuromotorie, Università degli Studi di Bologna, Bologna, Italy |
| Chiara La Morgia | IRCCS Istituto delle Scienze Neurologiche di Bologna, UOC Clinica Neurologica, Dipartimento di Scienze Biomediche e Neuromotorie, Università degli Studi di Bologna, Bologna, Italy |
| Marie-Bénédicte Rougier | Department of Ophthalmology, University Hospital of Bordeaux, Bordeaux, France |
| Étienne Bénard-Séguin | Queens University School of Medicine, Kingston, Ontario, Canada |
| Leonard B. Milea | University of California, Berkeley, Berkeley, California, USA |
| Selvakumar Ambika | Neuro-ophthalmology. Sankara Nethralaya-A unit of Medical Research Foundation, Chennai, India |
| Pedro L. Fonseca | Ophthalmology, Centro Hospitalar e Universitário de Coimbra (CHUC), Coimbra Institute for Biomedical Imaging and Translational Research (CIBIT), Faculty of Medicine University of Coimbra (FMUC), Coimbra, Portugal |
| Elisabeth Arnberg Wibroe | Department of Ophthalmology, Rigshospitalet, University of Copenhagen, Glostrup, Denmark |
| Nicolae Sanda | Department of Clinical Neurosciences, Geneva University School of Medicine, Geneva, Switzerland |
| Gabriele Thumann | Department of Clinical Neurosciences, Geneva University School of Medicine, Geneva, Switzerland |
| Christophe Chiquet | Department of Ophthalmology, University Hospital of Grenoble-Alpes, Grenoble-Alpes University, HP2 Laboratory, INSERM U1042, Grenoble, France |
| Hui Yang | Zhongshan Ophthalmic Center, Sun Yat-sen University, Guangzhou, P.R.China |
| Carmen K.M. Chan | Ophthalmology and Visual Sciences, Hong Kong Eye Hospital, The Chinese University of Hong Kong, Hong Kong Special Administrative Region, China |

| | |
|---|---|
| Carol Y. Cheung | Ophthalmology and Visual Sciences, Hong Kong Eye Hospital, The Chinese University of Hong Kong, Hong Kong Special Administrative Region, China |
| Janvier Ngoy Kilangalanga | Eye Department, Saint Joseph Hospital/Centre de Formation Ophtalmologique Pour l'Afrique Centrale, Kinshasa, Democratic Republic of the Congo |
| Makoto Nakamura | Division of Ophthalmology, Department of Surgery, Kobe University Graduate School of Medicine, Kobe, Japan |
| Fumio Takano | Division of Ophthalmology, Department of Surgery, Kobe University Graduate School of Medicine, Kobe, Japan |
| Neringa Jurkute | Moorfields Eye Hospital NHS Foundation Trust, London, UK<br>UCL Institute of Ophthalmology, University College London, London, UK |
| Patrick Yu-Wai-Man | Moorfields Eye Hospital NHS Foundation Trust, London, UK<br>UCL Institute of Ophthalmology, University College London, London, UK<br>Cambridge Eye Unit, Addenbrooke's Hospital, Cambridge University Hospitals, Cambridge, UK<br>Cambridge Centre for Brain Repair and MRC Mitochondrial Biology Unit, Department of Clinical Neurosciences, University of Cambridge, Cambridge, UK |
| Richard Kho | American Eye Center, Mandaluyong City, Manila, Philippines |
| John J. Chen | Department of Ophthalmology and Neurology, Mayo Clinic, Rochester, MN, USA |
| Jeong-Min Hwang | Ophthalmology Department, Seoul National University College of Medicine, Seoul National University Bundang Hospital, Republic of Korea |
| Dong Hyun Kim | Ophthalmology Department, Seoul National University College of Medicine, Seoul National University Bundang Hospital, Republic of Korea |
| Hee Kyung Yang | Ophthalmology Department, Seoul National University College of Medicine, Seoul National University Bundang Hospital, Republic of Korea |
| Jing Liang Loo | Neuro-Ophthalmology Department, Singapore National Eye Centre, Singapore |
| Reuben Chao Ming Foo | Neuro-Ophthalmology Department, Singapore National Eye Centre, Singapore |
| Shweta Singhal | Neuro-Ophthalmology Department, Singapore National Eye Centre, Singapore |
| Sharon Lee Choon Tow | Neuro-Ophthalmology Department, Singapore National Eye Centre, Singapore |
| Caroline Vasseneix | Visual Neuroscience Research Group, Singapore Eye Research Institute, Singapore National Eye Center, Singapore |
| Maxwell T. Finkelstein | Visual Neuroscience Research Group, Singapore Eye Research Institute, Singapore National Eye Center, Singapore |
| Luis J. Mejico | Departments of Neurology, Ophthalmology, SUNY Upstate Medical University, Syracuse, NY, USA |
| Masoud Aghsaei Fard | Farabi Eye Hospital, Tehran University of Medical Science, Tehran, Iran |
| Jonathan A. Micieli | Faculty of Medicine, University of Toronto, Toronto, Ontario, Canada<br>Department of Ophthalmology and Vision Sciences, University of Toronto, Toronto, Ontario, Canada<br>Kensington Vision and Research Centre, Toronto, Ontario, Canada<br>Department of Ophthalmology, St. Michael's Hospital and Toronto Western Hospital, Toronto, Ontario, Canada |

| | |
|---|---|
| Mukharram M. Bikbov | Ufa Eye Research Institute, Ufa, Russia |
